# Rapid SARS-CoV-2 Detection by Carbon Nanotube-Based Near-Infrared Nanosensors

**DOI:** 10.1101/2020.11.02.20223404

**Authors:** Rebecca L. Pinals, Francis Ledesma, Darwin Yang, Nicole Navarro, Sanghwa Jeong, John E. Pak, Lili Kuo, Yung-Chun Chuang, Yu-Wei Cheng, Hung-Yu Sun, Markita P. Landry

## Abstract

To effectively track and eliminate COVID-19, it is critical to develop tools for rapid and accessible diagnosis of actively infected individuals. Here, we introduce a single-walled carbon nanotube (SWCNT)-based optical sensing approach towards these ends. We construct a nanosensor based on SWCNTs noncovalently functionalized with ACE2, a host protein with high binding affinity for the SARS-CoV-2 spike protein. Presence of the SARS-CoV-2 spike protein elicits a robust, two-fold nanosensor fluorescence increase within 90 min of spike protein exposure. We characterize the nanosensor stability and sensing mechanism, and passivate the nanosensor to preserve sensing response in saliva and viral transport medium. We further demonstrate that these ACE2-SWCNT nanosensors retain sensing capacity in a surface-immobilized format, exhibiting a 73% fluorescence turn-on response within 5 s of exposure to 35 mg/L SARS-CoV-2 virus-like particles. Our data demonstrate that ACE2-SWCNT nanosensors can be developed into an optical tool for rapid SARS-CoV-2 detection.

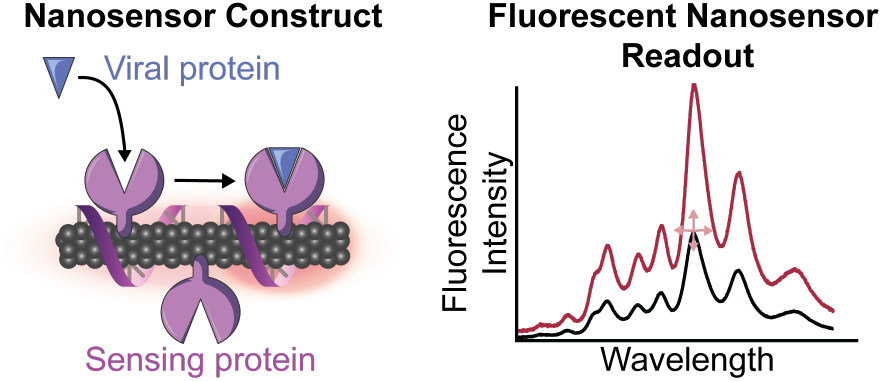

## Introduction

The World Health Organization deemed COVID-19 a global pandemic on March 11th, 2020. As of October 30th, SARS-CoV-2 has caused over 1 million deaths and infected almost 45 million people worldwide (*1*). It is estimated that over 70% of infected individuals under the age of 60 are asymptomatic, yet can still transmit the virus to others (*2*). Early estimates placed the basic reproductive number (R0) at 2.2, which represents the average number of people an infected person will spread the disease to (*3*). Taken together, these findings underscore the need for advancements in testing and containment efforts to end the pandemic. Pioneering examples towards such ends include Singapore’s successful COVID-19 containment plan (*4*) and the UC Berkeley Innovative Genomics Institute (IGI)’s pop-up testing center (*5*).

Current SARS-CoV-2 testing strategies can be grouped into two categories: molecular tests and serological tests. Molecular tests remain the status-quo for detecting active CoV-2 infections by detecting CoV-2 RNA in patient samples including sputum and nasal fluid. Molecular tests primarily use real-time reverse transcription polymerase chain reaction (RT-PCR), and are thus expensive ($5-10 per test), time-consuming (2-3 hours), and require laboratory processing (*6*–*9*). Yet, RT-PCR tests possess high sensitivity in identifying viral nucleic material, with the limit of detection (LOD) reported between 1-10 viral RNA copies necessary to produce a positive result (*6*). Serological tests detect the presence of IgG and IgM antibodies in a patient blood serum and provide important surveillance data of past viral infections, yet do not identify active cases. As such, detection of viral RNA by molecular tests is to-date the preferred testing mode to diagnose active CoV-2 cases. However, the complexity of the process necessitates the use of expensive equipment and trained personnel, limiting the testing capability in rural and lower income regions (*10*). Altogether, these factors amount to a large enough backlog in RT-PCR testing capabilities such that the United States is currently at 71% of its testing target to mitigate the spread of the virus as of October 4^th^ (*11*). Several non-PCR-based methods of viral RNA detection have been developed recently, harnessing techniques such as loop-mediated isothermal amplification (LAMP) (*12*), localized surface plasmon resonance (LSPR) (*13*), and CRISPR machinery (*14*) to avoid the expensive equipment required for the heating and cooling cycles of PCR. However, these techniques are not as sensitive as PCR and still require between 30 minutes and 1 hour of processing time per sample (*15*). Antigen testing has emerged with great potential for rapid diagnostics, possessing the key strength that active virus is detected. This contrasts with RT-PCR tests, which are merely detecting the presence of viral RNA and can consequently lead to cases of RT-PCR positivity in the absence of any viable virus (*16*). Such rapid tests are faster and cheaper yet possess lower sensitivity (*16*).

There has been a strong drive to find other viral testing targets and methodologies for simpler and faster diagnostics due to extended processing time and equipment restrictions associated with viral RNA detection. SARS-CoV-2 is a coronavirus, a family of viruses termed as such due to the halo or “corona” of proteins surrounding the virus in electron microscopy images. These outwardly protruding spike (S) proteins bind to the angiotensin-converting enzyme 2 (ACE2) receptor on the surface of human respiratory endothelial cells, facilitating viral entry (*17, 18*). The viral S protein is also the primary antigen that human monoclonal antibodies bind to in order to prevent host cell infusion and mark the virus for clearance (*19*). With approximately 100 S protein trimers present per SARS-CoV-2 virion, the S protein has become a prime target for live virus detection (*20*). For example, Seo et al. developed a field-effect transistor-based sensor by functionalizing graphene sheets with SARS-CoV-2 S protein antibody to detect SARS-CoV-2 at a LOD of 242 copies/mL in crude, nasopharyngeal swab clinical samples (*21*). Several promising nanotechnology-based sensors for SARS-CoV-2 detection have also emerged for both nucleic acid- and antigen-based detection and diagnosis of COVID-19, including platforms based on gold nanoparticles and quantum dots (*13, 22*–*25*). Such technologies will be crucial in working towards sensitive tests that do not rely on specialized equipment for signal readout and controlled laboratory environments for sample processing (*15*). Single-walled carbon nanotubes (SWCNTs) have shown much utility for biological analyte sensing (*26*–*28*). SWCNTs are intrinsically near-infrared (nIR) fluorescent and can be functionalized with various sensing moieties to develop stable biological sensors with rapid fluorescence-change readouts. Unlike conventional fluorophores, SWCNTs do not photobleach, giving rise to their potential long-term use (*27*). Importantly, the SWCNT near-infrared emission is minimally absorbed and scattered by biomolecules (*27*), providing a readout that can penetrate optically occluded patient samples, thus eliminating the need for sample purification and processing that limit the throughput of other testing modes. Furthermore, SWCNTs offer facile incorporation into portable form factors such as immobilization in paper or hydrogels (*29, 30*) with detection of the nIR SWCNT signal by a raspberry pi system, of similar form factor to a smartphone (*31*).

Herein we demonstrate the development and characterization of a SWCNT-based nanosensor capable of detecting SARS-CoV-2 via S-protein sensing. This nanosensor is based on the innate interactions of human host proteins with components of the SARS-CoV-2 virion, in conjunction with a SWCNT substrate to provide the optical readout. Specifically, host cell membrane protein ACE2 binds to the viral spike protein receptor-binding domain (S RBD) protruding from the virion surface. We constructed nanosensors by immobilizing ACE2 proteins on the surface of single-walled carbon nanotubes (SWCNTs) that serve to provide the nanosensor’s optical readout. Upon S protein binding to ACE2-functionalized SWCNTs, the change in dielectric environment surrounding the SWCNTs leads to a modulation in the nIR SWCNT fluorescence. This noncovalent modification strategy is advantageous towards retaining the intact SWCNT surface lattice that is necessary for fluorescence (*32*). We demonstrate that ACE2-functionalized SWCNT nanosensors can achieve a LOD of 9.5 nM S RBD, can be passivated for detection of S protein in saliva and viral transport medium, and can be imaged for rapid detection of S protein or virus-like CoV-2 virions within seconds.

## Results

### Nanosensor platform generation and characterization

To generate nanosensors, we first solubilized SWCNTs by probe-tip sonication with single-stranded DNA (ssDNA), (GT)_6_ (see **Methods**). Direct probe-tip sonication of ACE2 with pristine SWCNTs did not lead to a stable suspension and further raises the likelihood for disruption of the native ACE2 protein conformation, and hence loss of sensing ability for S RBD. The ssDNA sequence of (GT)_6_ was chosen based on high SWCNT suspension yield. The short ssDNA sequence length (12 nucleotides) was informed by previous work demonstrating that shorter ssDNA desorbs faster, and to a greater extent, from SWCNTs in the presence of proteins (*33, 34*). (GT)_6_-SWCNTs (2.5 mg/L final concentration) were incubated with ACE2 sensing protein (6.25 mg/L final concentration) in 0.1 M phosphate-buffered saline (PBS) solution to noncovalently passivate the SWCNT surface with protein, schematically represented in **Figure 1A** (*34*). This ratio of ACE2 sensing protein to SWCNT substrate was calculated to be approximately above the close-packing threshold to minimize protein surface-denaturation and colloidal aggregation, then this calculated value was experimentally optimized (see details in **SI** and **Figure S1, Figure S2**). ACE2 adsorption to (GT)_6_-SWCNTs manifested as a nearly instantaneous quenching of the SWCNT fluorescence, leveling off to −37% integrated-fluorescence fold change (*ΔF/F*_*0*_) within 5 min (**Figure 1B**). This fluorescence quenching exhibited excellent time stability over the course of 2 h (**Figure 1C**). Comparing the time-dependent quenching behavior of ACE2 with (GT)_6_-vs. (GT)_15_-SWCNTs affirmed the faster “leaving group” behavior of the shorter ssDNA, (GT)_6_, stabilizing within 10 minutes, as compared to (GT)_15_, requiring at least 60 minutes (**Figure S3**). Noncovalent ACE2 adsorption, as opposed to covalent modification, was confirmed by retention of SWCNT absorbance peaks representing various SWCNT chiralities’ electronic transitions (**Figure S4**). These results demonstrate that ACE2 adsorption on SWCNTs leads to increased nonradiative decay pathways, resulting in quenched nIR SWCNT fluorescence.

**Figure 1.**
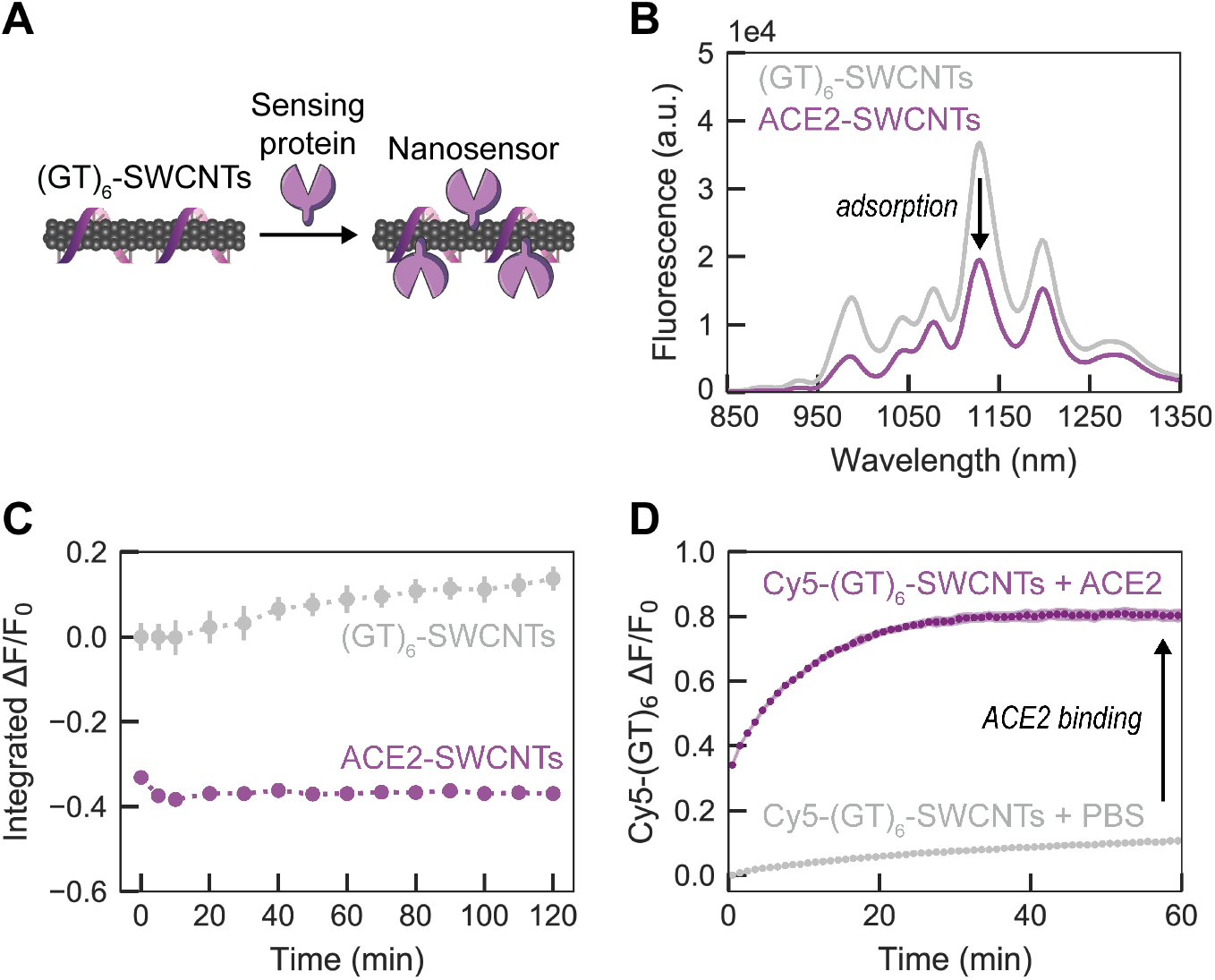
Adsorption of ACE2 sensing proteins to (GT)_6_-SWCNTs. **(A)** Schematic depiction of ACE2-SWCNT nanosensor formation, with sensing protein ACE2 adsorbing to (GT)_6_-SWCNTs. **(B)** ACE2-SWCNT complexation was observed as quenching of the intrinsic SWCNT near-infrared fluorescence following 1 h incubation of 6.25 mg/L ACE2 with 2.5 mg/L (GT)_6_-SWCNTs (final concentrations). **(C)** ACE2-SWCNT construct demonstrated time-stable quenched fluorescence. All fluorescence measurements were obtained with 721 nm laser excitation. **(D)** Adsorption of ACE2 on the SWCNT surface led to (GT)_6_ desorption, tracked by Cy5-labeled ssDNA following addition of 6.25 mg/L ACE2 with 2.5 mg/L Cy5-(GT)_6_-SWCNTs (final concentrations). The increase in Cy5-(GT)_6_ fluorescence from the initial quenched state on the SWCNT serves as a proxy for ACE2 adsorption. Shaded error bars represent standard error between experimental replicates (N = 3).

The affinity of ACE2 for the ssDNA-wrapped SWCNT surface was assessed by the corona exchange assay (*33*). For this assay, Cy5-labeled (GT)_6_ ssDNA was tracked as it desorbed from the SWCNT surface and thus de-quenched in the presence of ACE2. ACE2 displayed high affinity for the SWCNT surface, as ACE2 adsorption led to an 80.5% increase in Cy5 fluorescence, denoting free ssDNA, 1 h post addition of ACE2 (**Figure 1D**) in an ACE2 concentration-dependent manner (**Figure S5**). We further assessed the stability of the ACE2-SWCNT interface with a surfactant displacement assay, which confirmed strong and stable adsorption of ACE2 to the SWCNT (**Figure S6**) (*35, 36*). Taken together, these results suggest that ACE2 adsorbs to the SWCNT surface, displaces ssDNA originally on the SWCNT surface, and forms a stable ACE2-SWCNT conjugate that can be assessed for its response to S protein and for its utility as a CoV-2 nanosensor.

### Nanosensor response to SARS-CoV-2 spike protein

We analyzed the fluorescence response of ACE2-SWCNT nanosensors to the SARS-CoV-2 S RBD analyte, schematically represented in **Figure 2A**. Recognition of the CoV-2 S RBD by the nanosensor elicited a strong turn-on fluorescence response upon addition of 10 mg/L final concentration of CoV-2 S RBD to the nanosensor (formed by adsorbing 6.25 mg/L ACE2 to 2.5 mg/L (GT)_6_-SWCNTs) (**Figure 2B**). The normalized change in fluorescence of the 1130 nm SWCNT emission peak instantaneously increased to *ΔF/F*_*0*_ = 21.1%, reaching *ΔF/F*_*0*_ = 99.6% after 90 min (**Figure 2C**). This fluorescence modulation was verified to arise from the S RBD analyte itself rather than any impurities still remaining post-gel filtration chromatography (see **Methods**) by testing the filtrate of S RBD solution below a 3 kDa molecular weight cutoff centrifugal filter (**Figure S7**), which showed a negligible change in fluorescence above that of adding PBS. The concentration-dependent nanosensor response to S RBD (**Figure 2D**) gives rise to a 9.5 nM nanosensor LOD (see calculation in **SI**). Further, approximate values for the nanosensor kinetic parameters were determined by fitting the 90-minute nanosensor response to this analyte concentration series to the Hill Equation (cooperative binding model) (*28*). Here, the integrated-fluorescence fold change of the nanosensor was correlated to the concentration of the S RBD analyte as shown in **Figure 2E**, resulting in an equilibrium dissociation constant (*K*_*d*_) of 4.22 μM^-1^. These fit values represent conservative estimates for the nanosensor kinetic parameters by using the full integrated-fluorescence fold change. Moreover, this model implicates the assumption that the bulk analyte concentration remains constant (*i*.*e*. only a small fraction of total injected analyte is bound by the nanosensor). Importantly, the response of ACE2-SWCNT nanosensors for S RBD was confirmed by showing insignificant (GT)_6_ DNA desorption that does not scale with injected S RBD analyte concentration (**Figure S8A**). Furthermore, addition of S RBD to (GT)_6_-SWCNTs alone (without ACE2 sensing moiety) resulted in aggregation, as the ssDNA displaced from the SWCNT surface continues to increase linearly over 6 h (**Figure S8B**).

**Figure 2.**
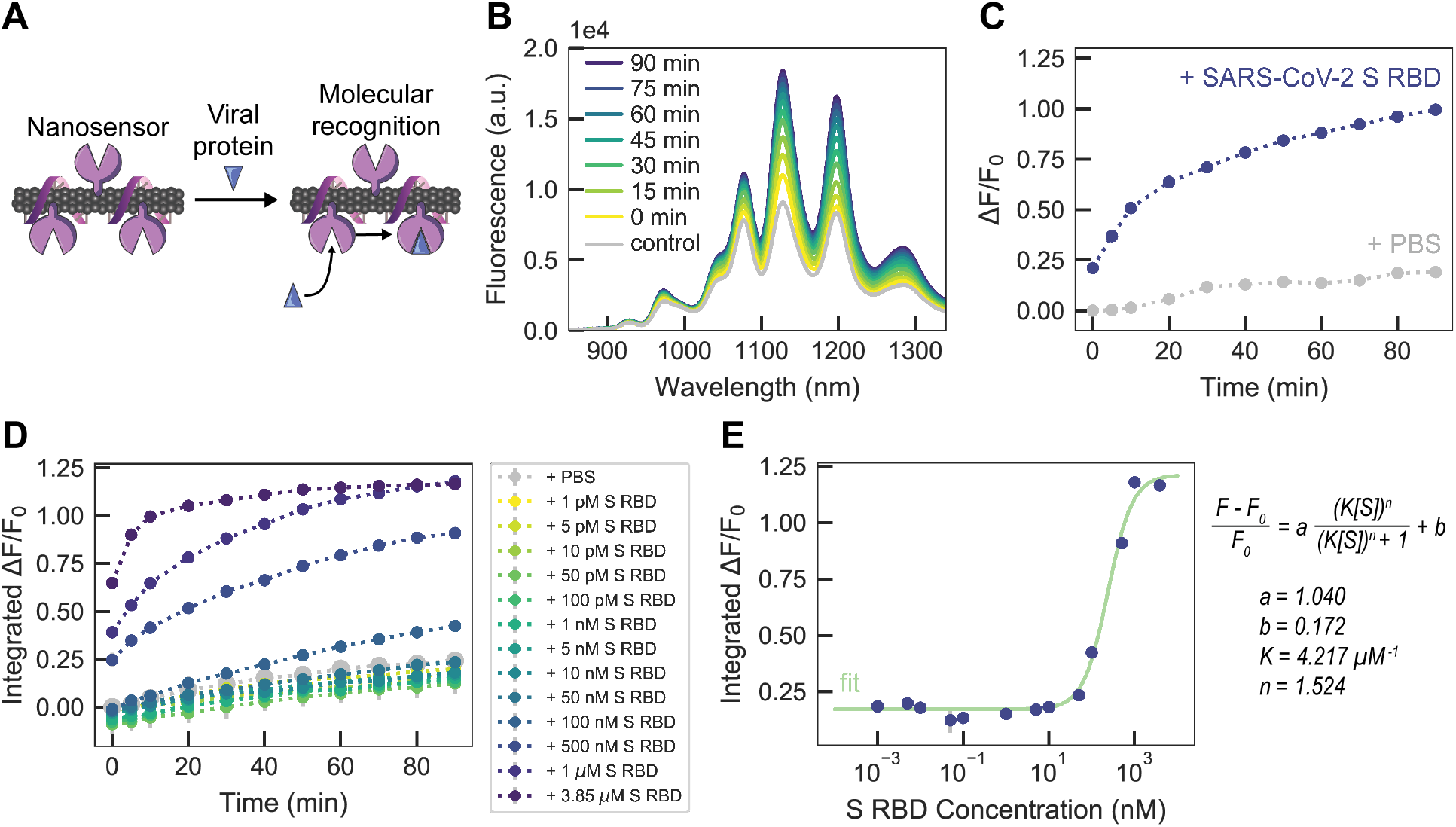
ACE2-SWCNT nanosensor response to SARS-CoV-2 spike protein receptor-binding domain (S RBD). **(A)** Schematic depiction of ACE2-SWCNT nanosensor interacting with viral protein S RBD. Addition of 10 mg/L S RBD (final concentration) to ACE2-SWCNTs (formed by 6.25 mg/L ACE2 and 2.5 mg/L (GT)_6_-SWCNTs) yielded a strong turn-on fluorescence response, as shown by **(B)** the full fluorescence spectrum and **(C)** the normalized change in fluorescence (*ΔF/F*_*0*_) of the 1130 nm SWCNT emission peak as a function of time, over 90 min. **(D)** Varying S RBD concentrations were injected into ACE2-SWCNTs and the integrated-fluorescence fold change (*ΔF/F*_*0*_) was monitored over 90 min. **(E)** Integrated *ΔF/F*_*0*_ values at time = 90 min for varying S RBD concentrations were fit to a cooperative binding model to quantify nanosensor kinetic parameters. All fluorescence measurements were obtained with 721 nm laser excitation.

Nanosensor colloidal stability was verified by demonstrating that the nanosensor response to S RBD persisted after centrifugation (16.1 krcf, 30 min; **Figure S9A**) and after overnight incubation at ambient conditions (**Figure S9B**). Repeating the surfactant displacement experiment with the ACE2-SWCNT nanosensor in the presence of S RBD showed that the bound receptor-ligand state further stabilized the nanosensor surface to surfactant perturbations (**Figure S6E-F**).

### Nanosensor analyte selectivity and bioenvironment robustness

We next investigated the selectivity of ACE2-SWCNT nanosensors to a panel of viral spike-like proteins. This viral analyte panel was composed of the SARS-CoV-2 S RBD in addition to the SARS-CoV-1 S RBD, MERS S RBD, and FLU hemagglutinin subunit (HA1). Serum albumin (HSA) was also included as a protein abundant in bioenvironments and in viral transport medium (VTM). Viral proteins were normalized on a mass basis (10 mg/L final concentration) to account for varying molecular weights. SARS-CoV-2 S RBD elicited the largest nanosensor response of *ΔF/F*_*0*_ = 99.6% at the 1130 nm SWCNT emission peak after 90 min (**Figure 3A**), followed by SARS-CoV-1 S RBD (*ΔF/F*_*0*_ = 88.4%). This cross-reactivity is expected, as ACE2 is also the cell membrane protein that binds to CoV-1 S RBD, although at ∼10 to 20-fold lower affinity (*37, 38*). MERS and FLU spike-like proteins naturally interact with different cell membrane receptors, accounting for this lower magnitude fluorescence response with our ACE2-SWCNT nanosensors.

**Figure 3.**
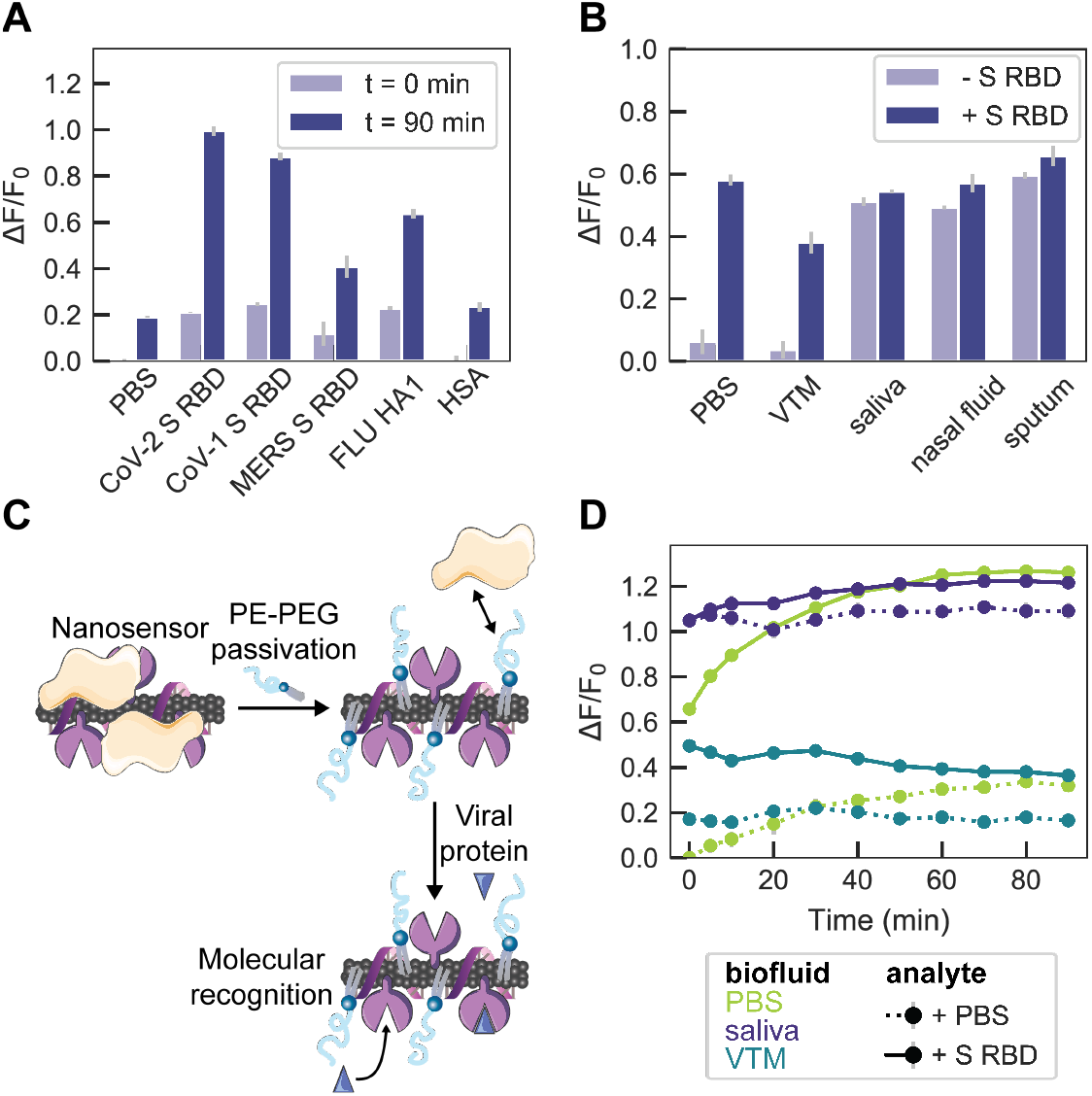
ACE2-SWCNT nanosensor selectivity and sensitivity in biofluid environments. **(A)** Normalized change in fluorescence (*ΔF/F*_*0*_) of the 1130 nm SWCNT emission peak for the ACE2-SWCNT nanosensor 0 min and 90 min after exposure to 10 mg/L of viral protein panel: SARS-CoV-2 spike receptor-binding domain (S RBD), SARS-CoV-1 S RBD, MERS S RBD, and FLU hemagglutinin subunit (HA1). **(B)** ACE2-SWCNT nanosensor response 90 min after exposure to 1 μM S RBD in the presence of 1% relevant biofluids: viral transport medium (VTM), saliva, nasal fluid, and sputum (treated with sputasol). **(C)** Schematic depiction of nanosensor biofouling with proteins present in relevant biofluids, mitigated upon passivation with phosphatidylethanolamine phospholipid with a 5000 Da PEG chain (PE-PEG). **(D)** Response of PE-PEG passivated nanosensor to 500 nM S RBD in the presence of PBS, 10% VTM, or 1% saliva. Surface passivation with a hydrophilic polymer improved the nanosensor response that was otherwise greatly attenuated, as shown in **(B)**. All fluorescence measurements were obtained with 721 nm laser excitation.

Nanosensor compatibility in biofluids was assessed by testing the nanosensor response to CoV-2 S RBD in 1% relevant biological fluids, including viral transport medium (VTM), human saliva, human nasal fluid, and human sputum (treated with sputasol) (biofluid details in **Table S1**). Although the nanosensor response was maintained in PBS and VTM, the response was diminished in the other biofluids, with a *ΔF/F*_*0*_ = 3.2% in saliva, 7.8% in nasal fluid, and 6.3% in sputum (**Figure 3B**). The attenuation of nanosensor response seems to arise from biofluid protein adsorption to the nanosensor surface that raises the baseline fluorescence and obfuscates viral analyte interaction, whereby the nanosensor fluorescence in the biofluids alone is stable with an increased baseline fluorescence (**Figure S9C**).

To mitigate the unfavorable effects of biofouling that lead to this diminished nanosensor response, we pursued a passivation strategy involving phosphatidylethanolamine phospholipid with a 5000 Da PEG chain attached to the head group (PE-PEG), schematically represented in **Figure 3C** (*39*). The PE-PEG passivated nanosensor response to 500 nM CoV-2 S RBD was *ΔF/F*_*0*_ = 19.9% in 10% VTM (otherwise absent without passivation) and *ΔF/F*_*0*_ = 12.4% in 1% saliva (otherwise 3.2% without passivation), suggesting PE-PEG nanosensor passivation enables partial mitigation of nanosensor biofouling.

### Immobilized nanosensor response to spike protein and virus-like particles

We next translated the nanosensors from in-solution sensing to a surface-immobilized format for imaging. ACE2-SWCNTs (formed by adsorbing 12.5 mg/L ACE2 to 5 mg/L (GT)_6_-SWCNTs) were immobilized on a glass bottom microwell dish and imaged with a 100x oil immersion objective (**Figure 4**). Addition of PBS at 60 s did not cause a change in fluorescence signal, as anticipated based on solution-based nanosensor control experiments (**Figure 2**). Upon injection of 2 μM (final concentration) S RBD, the average integrated-fluorescence intensity change was *ΔF/F*_*0*_ = 65.1% within 5 s (**Figure 4A-C**). This experiment was repeated using virus-like particles (VLPs), which are formed by co-expressing all four SARS-CoV-2 structural proteins (spike, membrane, nucleocapsid, and envelope proteins). Addition of 10% sucrose (the VLP buffer) at 60 s slightly increased the baseline fluorescence. Injection of 35 mg/L VLPs increased the average integrated-fluorescence intensity by *ΔF/F*_*0*_ = 72.8% within 5 s (**Figure 4D-F**). This concentration of VLPs corresponds to approximately 17 nM S RBD. To evaluate the specificity of the observed nanosensor response, we then tested VLPs produced with and without S protein co-expressed. We found that the immobilized nanosensor exhibited a response of *ΔF/F*_*0*_ = 19.4% for the VLPs without S protein, compared to a response of *ΔF/F*_*0*_ *=* 70.7% for the VLPs with S protein (**Figure S10**).

**Figure 4.**
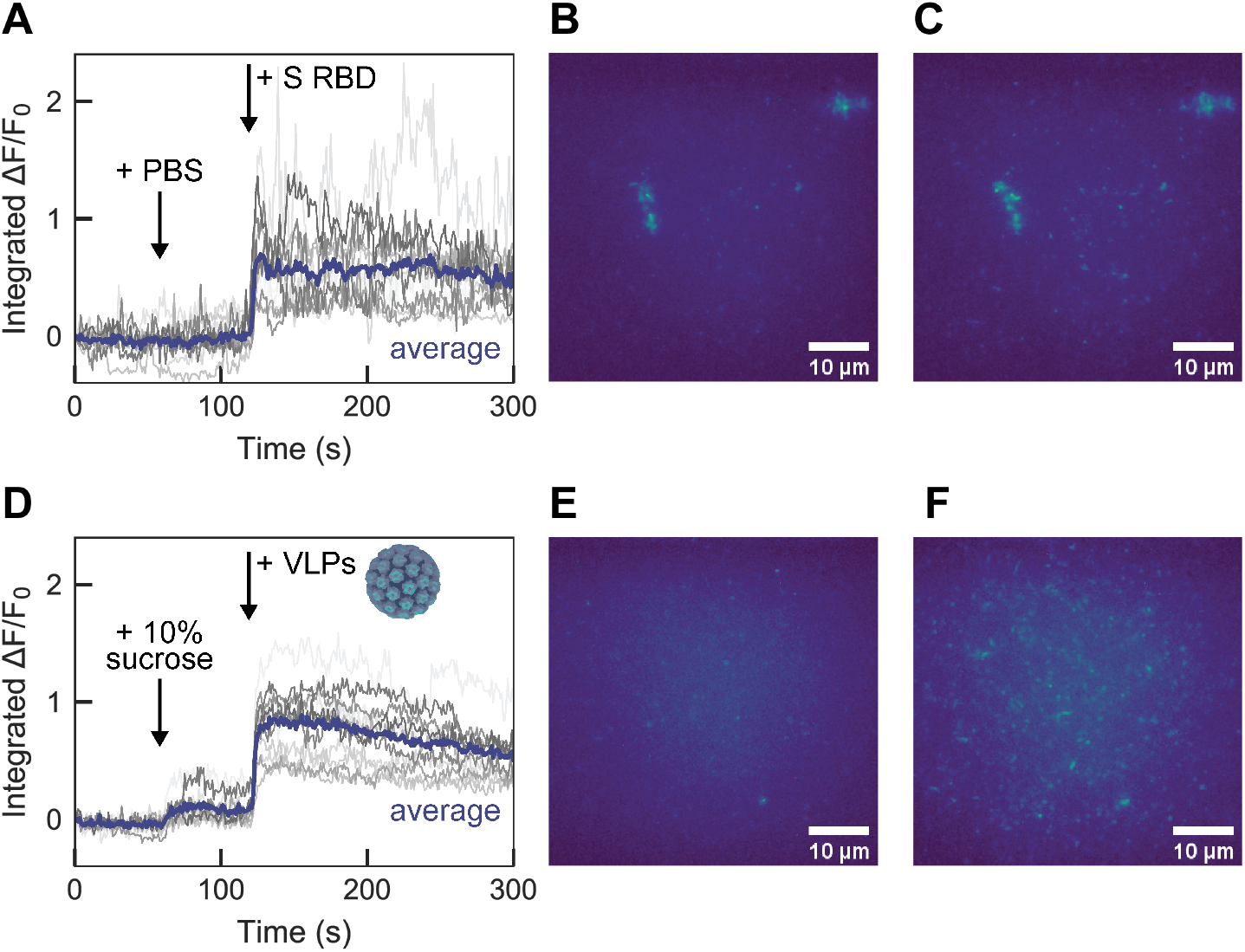
Surface-immobilized ACE2-SWCNT nanosensor response to SARS-CoV-2 spike protein receptor-binding domain (S RBD) and virus-like particles (VLPs). Single-molecule microscopy traces of ACE2-SWCNTs (formed by 12.5 mg/L ACE2 and 5 mg/L (GT)_6_-SWCNTs) immobilized on a glass microwell dish exhibited a fluorescence response to both S RBD and VLPs, for single regions of interest (gray; 12 total per image) and the average intensity (purple). **(A-C)** Addition of PBS at 60 s caused no change in fluorescence, as expected, and addition of 2 μM S RBD (final concentration) at 120 s yielded a turn-on fluorescence response, as shown by **(A)** the integrated-fluorescence fold change (*ΔF/F*_*0*_) over 5 min and entire field-of-view at **(B)** time = 0 s and **(C)** time = 125 s. **(D-F)** Addition of 10% sucrose buffer (to match VLP buffer) at 60 s caused a slight increase in fluorescence and addition of 35 mg/L VLPs (final concentration) at 120 s yielded a turn-on fluorescence response, as shown by **(D)** the integrated-fluorescence fold change (*ΔF/F*_*0*_) over 5 min and entire field-of-view at **(E)** time = 0 s and **(F)** time = 125 s. All fluorescence images were obtained with 721 nm laser excitation and a 100x oil immersion objective.

## Discussion

Early and frequent testing is key to trace and control the spread of COVID-19. However, current diagnostics suffer from insufficient supply and throughput, where our reliance on tests with long turnaround times leads to delays in patients receiving test results. The early detection of active SARS-CoV-2 infections could limit high density congregations that promote viral spread, thus a technology capable of rapidly detecting active infections in crude biofluids is needed. Accordingly, we have developed a nanosensor to detect SARS-CoV-2, with potential to operate in crude biofluids. This nanosensor can detect virus-like particles to an S protein RBD equivalent of 17 nM with a *ΔF/F*_*0*_ = 73% nanosensor response within 5 s (**Figure 4D-F**). Ultimately, these nanosensors can be formulated into a point-of-care device for rapid diagnosis of individuals actively infected with SARS-CoV-2, using accessible equipment from a different supply chain than that of current testing modes.

Our nanosensor concept exploits the innate binding capabilities of host proteins with virion components, coupled to a SWCNT substrate for signal transduction. Specifically, we employed noncovalent modification of nIR-fluorescent SWCNTs with ACE2 membrane receptor protein. SWCNTs are optimal signal transduction elements for biological sensing due to their photostability and emission in the nIR over which crude biological fluids are maximally transparent. This nIR fluorescence enables nanosensor readout through the highly scattering biological fluids in which the nanosensors are intended for use: saliva, nasal fluid, sputum, and/or blood. We employed ACE2 as the sensing protein due to its recognition abilities towards coronavirus components, specifically the spike proteins on the CoV-2 surface. ACE2 was noncovalently adsorbed to the SWCNT surface and, in the presence of the viral S protein analyte, binding elicited a change in the local dielectric environment of the SWCNT substrate and resulted in a modulation of the intrinsic SWCNT fluorescence. We confirmed ACE2 adsorption to SWCNTs by employing corona exchange and surfactant displacement assays, which confirmed stable binding of ACE2 to SWCNT, and subsequently confirmed S RBD binding to the ACE2-SWCNT nanosensor. The resulting ACE2-SWCNT nanosensor constructs displayed long-term time stability and excellent colloidal stability and retained its binding specificity for CoV-2 S RBD in the surface-immobilized state. Nanosensors exhibited a 100% turn-on response in fluorescence upon addition of 1 μM CoV-2 S RBD, with response scaling as a function of concentration. Fitting to a cooperative binding model gave rise to kinetic parameter estimates to quantify nanosensor performance. Although the solution-phase LOD of 9.5 nM remains below that of conventional testing, the surface-immobilized nanosensor can achieve a *ΔF/F*_*0*_ = 73% within 5 s of exposure to a 17 nM S protein equivalent of VLPs, as a synthetic mimic of the full SARS-CoV-2 virion. When this experiment was repeated for VLPs without S protein, the response was reduced to *ΔF/F*_*0*_ = 19%. This result supports our hypothesis that molecular recognition is enabled by the specific ACE2-S protein interaction. Furthermore, solution-phase nanosensor passivation with a hydrophilic polymer (PEG) attached to phospholipids (PE) provided some improvement of nanosensor response to S protein in 10% VTM and 1% saliva. However, additional strategies must be pursued to further mitigate biofouling while retaining the fluorescence response.

Taken together, our data show that SWCNT-based nanosensors noncovalently functionalized with the human ACE2 receptor can detect the SARS-CoV-2 S protein in relevant patient biofluids and can be immobilized and imaged on microfluidic surfaces. Though less sensitive than PCR-based testing, the rapid 5 s nanosensor response in the surface-immobilized state towards SARS-CoV-2 VLPs has distinct advantages in enabling a rapid response for on-site testing, and has the potential to be used to detect SARS-CoV-2 without patient biofluid sample processing and purification. As such, this technology is amenable to incorporation into a point-of-care device and can serve as an intermediate triage step to rapidly separate the ever-growing backlog of patient samples into definite negatives and potential positives for further testing.

## Materials and Methods

### Synthesis of ssDNA-SWCNTs

Suspensions of single-walled carbon nanotube (SWCNTs) with single-stranded DNA (ssDNA) were prepared by mixing 0.2 mg of mixed-chirality SWCNTs (small diameter HiPco™ SWCNTs, NanoIntegris) with 250 µM of ssDNA (custom ssDNA oligos with standard desalting, Integrated DNA Technologies, Inc.) in 1 mL, 0.1 M phosphate-buffered saline (PBS). This mixture was bath sonicated for 30 min (Branson Ultrasonic 1800) then probe-tip sonicated for 10 min in an ice bath (3 mm probe tip at 50% amplitude, 5-6 W, Cole-Parmer Ultrasonic Processor). Suspensions were centrifuged to pellet insoluble SWCNT bundles and contaminants (16.1 krcf, 90 min). Supernatant was collected and ssDNA-SWCNT concentration was calculated with measured sample absorbance at 632 nm (NanoDrop One, Thermo Scientific) and the empirical extinction coefficient ε_632nm_=0.036 L mg^-1^ cm^-1^.(*26*) ssDNA-SWCNTs were stored at 4°C and diluted to a working concentration of 10 mg L^-1^ in 0.1 M PBS at ambient temperature ≥ 2 h prior to use.

### Preparation of proteins and biofluids

Proteins were sourced and reconstituted as listed in **Table S1**. All viral protein analytes were purified with desalting columns to remove impurities (Zeba Spin Desalting Columns, 0.5 mL with 7 kDa MWCO, Thermo Fisher Scientific) by washing with 0.1 M PBS three times (1500 rcf for 1 min), centrifuging with sample (1500 rcf for 2 min), and retaining sample in flow-through solution. Resulting protein concentration was measured with the Qubit Protein Assay (Thermo Fisher Scientific) and proteins were diluted in 0.1 M PBS to 10x the intended final analyte concentration.

Biofluids were prepared by centrifuging to remove any large contaminants (1000 rcf for 5 min) then diluting in 0.1 M PBS to 10x the intended final concentration. Sputasol was used to liquify sputum prior to use, used according to manufacturer’s instructions.

### Synthesis of ACE2-ssDNA-SWCNT nanosensors

Nanosensors were made by preparing solutions of 10 mg/L (GT)_6_-SWCNTs and 25 mg/L ACE2, mixing in equal volumes, incubating for 30 min, diluting by half with 0.1 M PBS, and incubating for an additional 30 min. Final concentrations of components are thus 2.5 mg/L (GT)_6_-SWCNTs and 6.25 mg/L ACE2. For the stability test of nIR fluorescence as a function of time, (GT)_6_-SWCNTs and ACE2 were injected together to these final concentrations and measured immediately.

For passivation of nanosensors with phospholipid-PEG, the protocol was slightly modified to incorporate first adsorption of the sensing protein (ACE2) then passivation of remaining exposed SWCNT surface by phospholipid-PEG (saturated 16:0 phosphatidylethanolamine-PEG 5000 Da, or PE-PEG). Passivated nanosensors were made by preparing solutions of 10 mg/L (GT)_6_-SWCNTs and 25 mg/L ACE2, mixing in equal volumes, incubating for 15 min, adding PE-PEG to a final concentration of 2.5 mg/L, diluting by half with 0.1 M PBS, bath sonicating for 15 min, and incubating for an additional 30 min.

### Preparation of SARS-CoV-2 S RBD analyte

Plasmid encoding for SARS-CoV-2 S RBD (*40*) was transiently transfected into suspension Expi293 cells at 0.5-1 L scale. Three days after transfection, cell culture supernatants were clarified and purified by Ni-NTA affinity chromatography as previously described (*41*) and the eluted protein was dialyzed extensively against PBS prior to storage at −80C.

### Synthesis and purification of SARS-CoV-2 VLPs

To prepare the SARS-CoV-2 VLPs, two plasmids pcDNA3.1-Spike and pIRES2-MNE were synthesized based on the sequence of the Wuhan-Hu-1 strain (GenBank: MN908947.3). The spike protein was stabilized with the furin cleavage (residues 682-685) abrogated and the consecutive residue 986 and 987 substituted with prolines (*38, 42*). The VLPs were synthesized by co-transfecting HEK293 or HEK293T cells with plasmids using HyFect transfection reagent (Leadgene Biomedical Inc., Taiwan) or JetOptimus (Polyplus-transfection, USA). To generate VLPs without S protein, cells were transfected with pIRES2-MNE only. The harvested supernatant was first concentrated with a 100 kDa MWCO centrifugal filter (Pall Corporation) then laid over discontinuous 20%-60% sucrose or Opti-prep (BioVision Inc.) gradients followed with centrifugation at 30,000 rpm for 4 hours. Purified VLPs were resuspended in PBS pH 7.4 and frozen at −80°C for storage.

### Nanosensor optical characterization and analyte screening

Fluorescence was measured with an inverted Zeiss microscope (Axio Observer.D1, 10x objective) coupled to a Princeton Instruments spectrometer (SCT 320) and liquid nitrogen-cooled Princeton Instruments InGaAs detector (PyLoN-IR). Samples were excited with a triggered 721 nm laser (OptoEngine LLC) and emission was collected in the 800 – 1400 nm wavelength range, with samples in a polypropylene 384 well-plate format (30 μL total sample volume; Greiner Bio-One microplate).

For nIR fluorescence screens, 27 µL of nanosensor was added per well and 3 µL of 10x-concentrated viral protein analytes in 0.1 M PBS (or buffer alone) were injected per well using a microchannel pipette (in triplicate), with brief mixing by pipetting. The plate was sealed with an adhesive seal (Bio-Rad) and spun down for 15 sec with a benchtop well plate centrifuge. Fluorescence spectra were recorded at time points of 0 min, 5 min, 10 min, and every subsequent 10 min until the max time point.

For surfactant stability tests, the screening protocol was modified as follows: 24 µL of nanosensor, 3 µL of 2.5 w/v% sodium cholate (SC), then 3 µL of 10x-concentrated viral protein analytes in 0.1 M PBS (or buffer alone) were added per well. Wavelength shifts were calculated by translating fluorescence spectra in 1 nm wavelength increments such that the correlation coefficient was maximized with respect to the reference state. Data processing in this manner captures the full spectrum shifting behavior.

Absorbance was measured by UV-VIS-nIR spectrophotometer (Shimadzu UV-3600 Plus) with samples in a 50 μL volume, black-sided quartz cuvette (Thorlabs, Inc.).

For surface-immobilized nanosensor experiments, ACE2-SWCNTs were immobilized on MatTek glass bottom microwell dishes (35 mm petri dish with 10 mm microwell) as follows: the dish was washed twice with 150 μL 0.1 M PBS, 100 μL of nanosensor (formed by 12.5 mg/L ACE2 with 5 mg/L (GT)_6_-SWCNT pre-incubated for 1 h) was added and incubated for 20 min, nanosensor solution was removed, and the dish was washed twice again with 150 μL 0.1 M PBS. Surface-immobilized nanosensors were imaged on an epifluorescence microscope (100x oil immersion objective) with an excitation of 721 nm and a Ninox VIS-SWIR 640 camera (Raptor). For each imaging experiment, 120 μL 0.1 M PBS was added prior to recording and the z-plane was refocused. Images were collected with a 950 ms exposure time and 1000 ms repeat cycle over 5 min. 15 μL buffer was added at frame 60 and 15 μL analyte was added at frame 120. Images were processed in ImageJ by applying a median filter (0.5-pixel radius) and rolling ball background subtraction (300-pixel radius), then using the ROI analyzer tool (Multi Measure).

### Corona exchange assay

Corona dynamic studies were done as described previously (*33*). Briefly, the same ssDNA-SWCNT suspension protocol was employed, instead using fluorophore-labeled ssDNA-Cy5 (3’ Cy5-labeled custom ssDNA oligos with HPLC purification, Integrated DNA Technologies, Inc.). Fluorescently labeled ssDNA was tracked and the displacement of ssDNA from the SWCNT surface (monitored as in increase in Cy5 fluorescence) was used as a proxy for protein adsorption. To assess (GT)_6_-Cy5 desorption from SWCNTs in the presence of ACE2, 25 µL of 12.5 mg L^-1^ ACE2 was added to 25 µL of 5 mg L^-1^ (GT)_6_-Cy5-SWCNTs. To assess (GT)_6_-Cy5 desorption from nanosensors in the presence of S RBD, 5 µL of 10x-concentrated S RBD was injected into 45 µL of ACE2-(GT)_6_-Cy5-SWCNTs. Solutions were added via microchannel pipette into a 96-well PCR plate (Bio-Rad) and mixed by pipetting. The plate was sealed with an optically transparent adhesive seal (Bio-Rad) and briefly spun down on a benchtop centrifuge. Fluorescence time series readings were measured in a Bio-Rad CFX96 Real Time qPCR System by scanning the Cy5 channel every 30 s at 22.5°C.

## Supporting information

Manuscript file

## Data Availability

All data needed to evaluate the conclusions in the paper are present in the paper and/or the Supplementary Materials.

## Acknowledgements

We acknowledge support of the IGI LGR ERA and Citris/Banatao Seed Funding. We acknowledge support of an NIH NIDA CEBRA award # R21DA044010 (to M.P.L.), a Burroughs Wellcome Fund Career Award at the Scientific Interface (CASI) (to M.P.L.), the Simons Foundation (to M.P.L.), a Stanley Fahn PDF Junior Faculty Grant with Award # PF-JFA-1760 (to M.P.L.), a Beckman Foundation Young Investigator Award (to M.P.L.), and a DARPA Young Investigator Award (to M.P.L.). M.P.L. is a Chan Zuckerberg Biohub investigator. R.L.P., F.L., and D.Y. acknowledge the support of NSF Graduate Research Fellowships (NSF DGE 1752814).

