## Supplementary material for "Rapid SARS-CoV-2 Detection by Carbon Nanotube-Based Near-Infrared Nanosensors": Manuscript file

The ratio of ACE2 to $(GT)_6$-SWCNTs was chosen based on a protein footprint estimation that 28.6 ACE2 dimers fit per SWCNT in the close-packed limit (using ACE2 dimer dimensions as determined by cryo-EM (*1*)). This calculation translates to a mass ratio of 2.36 ACE2:SWCNT. The actual mass ratio of 2.5 ACE2:SWCNT was chosen to be just above this theoretical close-packed limit in an attempt to minimize protein spreading that arises from a large excess of nanoparticle surface available for proteins. Extending to twice this mass ratio produces more quenching in the SWCNT fluorescence (**Figure S1**), yet reduced sensing ability (**Figure S2**) and colloidal stability. Thus, the mass ratio of 2.5 ACE2:SWCNT was experimentally determined to be best suited for S RBD sensing (**Figure S2**).

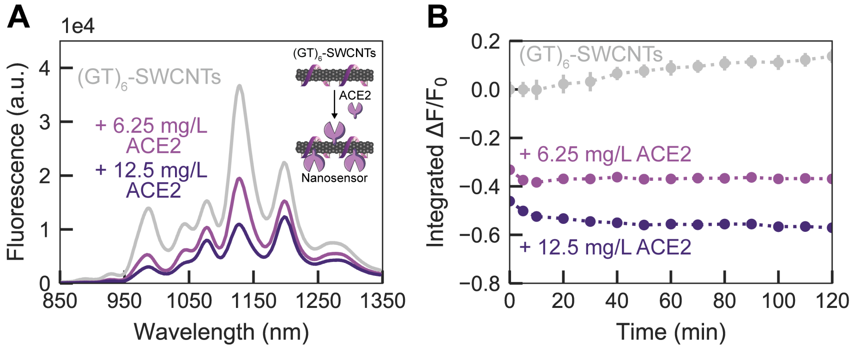


**Figure S1**. **Adsorption of different ratios of ACE2 on $(GT)_6$-SWCNTs.** ACE2-SWCNT complexation quenched intrinsic SWCNT near-infrared fluorescence, shown by **(A)** the full fluorescence spectrum after 1 h incubation of 6.25 mg/L or 12.5 mg/L ACE2 with 2.5 mg/L $(GT)_6$-SWCNTs (final concentrations) and **(B)** the integrated-fluorescence change as a function of time over 2 h. Both ratios exhibit stable quenched fluorescence, however, the lower ratio of protein to SWCNT exhibited better colloidal stability.

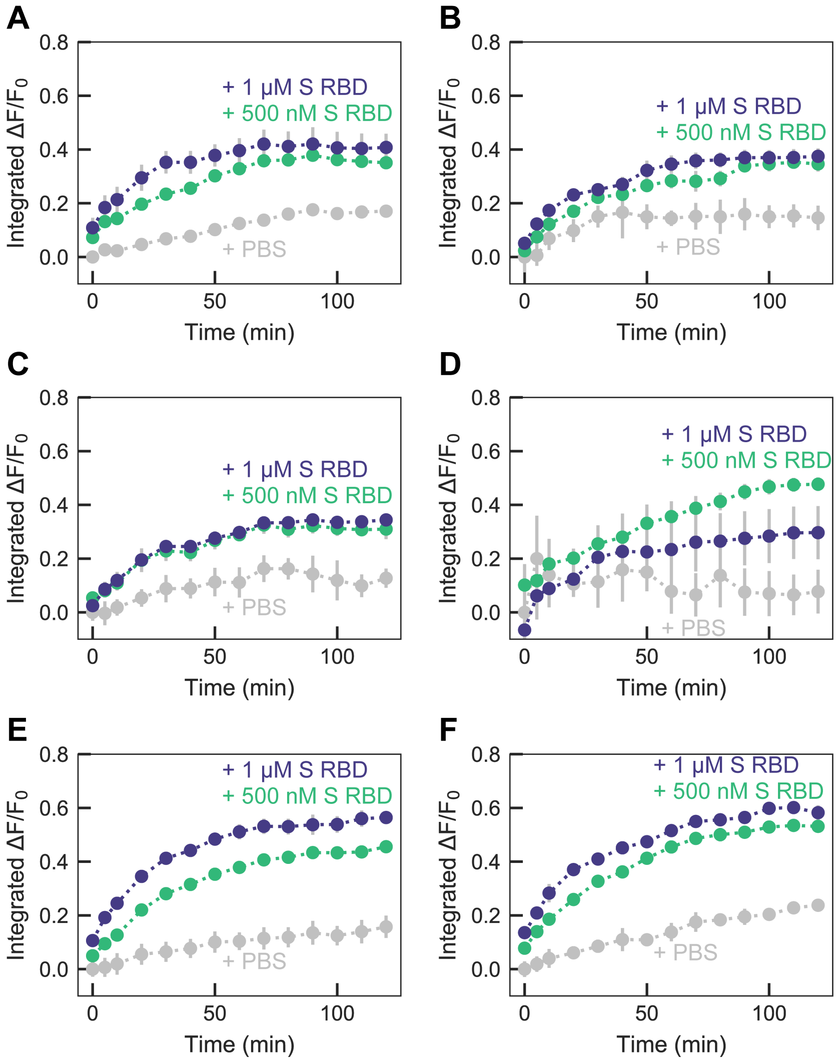


**Figure S2. Response of different ratios ACE2:(GT)$_6$-SWCNTs to S RBD.** Integrated-fluorescence fold change as a function of time over 120 min upon addition of PBS, 500 nM S RBD, or 1 µM S RBD to nanosensor formed by **(A)** 6.25 mg/L ACE2 and 2.5 mg/L (GT)$_6$-SWCNTs incubated 1 h, **(B)** 12.5 mg/L ACE2 and 2.5 mg/L (GT)$_6$-SWCNTs incubated 1 h, **(C)** 6.25 mg/L ACE2 and 2.5 mg/L (GT)$_6$-SWCNTs incubated 3 h, **(D)** 12.5 mg/L ACE2 and 2.5 mg/L (GT)$_6$-SWCNTs incubated 3 h, **(E)** 6.25 mg/L ACE2 and 2.5 mg/L (GT)$_6$-SWCNTs incubated 30 min, diluted by half in PBS, and incubated an additional 30 min, and **(F)** 12.5 mg/L ACE2 and 2.5 mg/L (GT)$_6$-SWCNTs incubated 30 min, diluted by half in PBS, and incubated an additional 30 min.

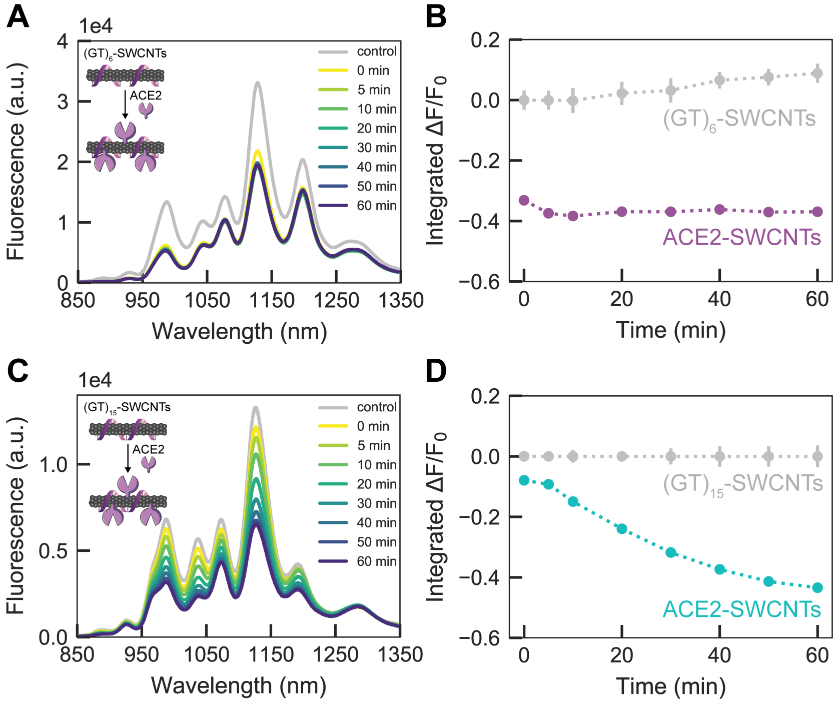


**Figure S3. Adsorption of ACE2 to $(GT)_6$- vs. $(GT)_{15}$-SWCNTs.** ACE2-SWCNT complexation rapidly quenched intrinsic SWCNT near-infrared fluorescence, shown by **(A)** the full fluorescence spectrum and **(B)** the integrated-fluorescence fold change as a function of time over 1 h, upon incubation of 6.25 mg/L ACE2 with 2.5 mg/L $(GT)_6$-SWCNTs (final concentrations). ACE2-SWCNT complexation quenched intrinsic SWCNT near-infrared fluorescence at a slower rate, shown by **(C)** the full fluorescence spectrum and **(D)** the integrated-fluorescence fold change as a function of time over 1 h, upon incubation of 6.25 mg/L ACE2 with 2.5 mg/L $(GT)_{15}$-SWCNTs (final concentrations).

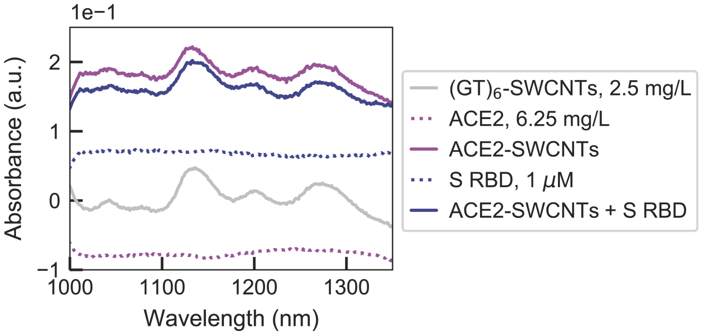


**Figure S4**. **Absorbance of $(GT)_6$-SWCNTs with ACE2 sensing protein and S RBD analyte.** Retention of near-infrared absorbance peaks arising from SWCNT in the case of ACE2 adsorption and S RBD analyte binding confirms the noncovalent passivation, rather than covalent modification, of the SWCNT surface.

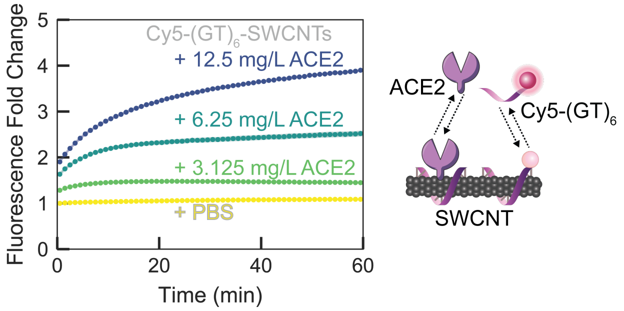


**Figure S5. Displacement of Cy5-$(GT)_6$ ssDNA from SWCNT as a function of passivating ACE2 concentration.** Adsorption of ACE2 on the SWCNT surface led to $(GT)_6$ desorption, tracked by Cy5-labeled ssDNA, in a concentration-dependent manner upon addition of varying ACE2 concentrations with 2.5 mg/L Cy5-$(GT)_6$-SWCNTs (final concentrations). The increase in Cy5-$(GT)_6$ fluorescence from the initial quenched state on the SWCNT serves as a proxy for ACE2 adsorption. Shaded error bars represent standard error between experimental replicates (N = 3).

To test the stability of the ACE2-functionalized SWCNT construct, we implemented a solvatochromic shift assay (*2*, *3*). This methodology is based upon addition of surfactant (sodium cholate, SC) that coats any solvent-exposed SWCNT surface and can displace low-affinity molecules from the SWCNT surface. SC adsorption causes exclusion of water from the SWCNT surface, producing a large increase in fluorescence and solvatochromic shift to lower emission wavelengths. Prior to ACE2 incubation, SC elicited both a large blue-shift (-16 nm) and sizeable increase in fluorescence (70.2%) for $(GT)_6$-SWCNTs alone (**Figure S6A-B**). However, upon passivation with ACE2, these spectral changes were reduced to -10 nm and 10.3%, respectively (**Figure S6C-D**), suggesting a mechanism in which ACE2 adsorbs to the SWCNT surface and causes $(GT)_6$ ssDNA desorption. Moreover, we show that addition of S RBD analyte to the ACE2-SWCNT complex further stabilizes the nanosensor, whereby addition of SC produced a brief blue-shift of -6 nm yet a complete return to baseline by 70 min, and similarly, a minimal change in fluorescence of -3.9% (**Figure S6E-F**).

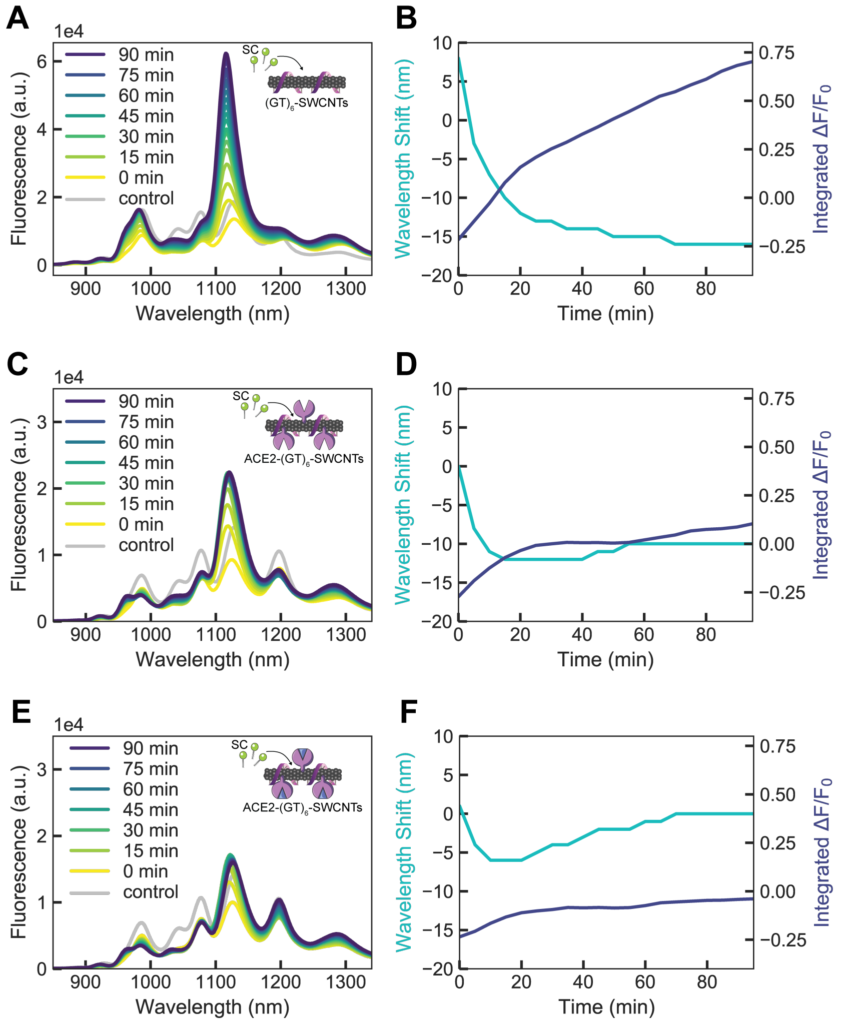


**Figure S6. Surfactant displacement experiment to probe the stability of (GT)$_6$-SWCNTs with ACE2 sensing protein and S RBD analyte.** Full fluorescence spectrum (left) and time-dependent wavelength shift and integrated-fluorescence fold change (right) for **(A-B)** 2.5 mg/L (GT)$_6$-SWCNTs alone, **(C-D)** ACE2-SWCNT nanosensors (formed by 6.25 mg/L ACE2 and 2.5 mg/L (GT)$_6$-SWCNTs), and **(E-F)** ACE2-SWCNT nanosensors with 500 nM S RBD, each upon addition of 0.25 w/v% sodium cholate (SC, final concentration). Decreasing blue-shift and fluorescence fold change due to ACE2 implies ACE2 covers and stabilizes the SWCNT surface, with further stabilization upon addition of S RBD analyte.

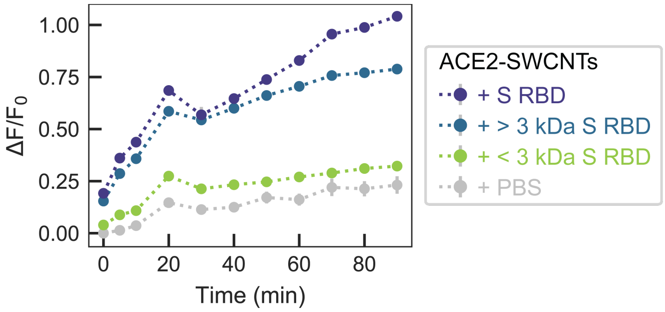


**Figure S7. ACE2-SWCNT nanosensor response to controls**. Addition of 500 nM S RBD (final concentration) to ACE2-SWCNTs (formed by 6.25 mg/L ACE2 and 2.5 mg/L $(GT)_6$-SWCNTs) yielded a significant turn-on fluorescence response. This response is maintained for S RBD > 3 kDa MWCO centrifugal filter and absent for S RBD < 3 kDa.

The limit of detection (*LOD*) of the ACE2-SWNT nanosensor for S RBD analyte is defined as the lowest analyte concentration likely to be reliably distinguished from the limit of blank (*LOB*) (*4*). The *LOB* is the highest apparent analyte concentration expected to be found when replicates of a blank sample containing only buffer (no analyte) are tested:

$$LOB = mean_{blank} + 1.645\ SE_{blank}$$

Where the mean and standard error (*SE*) are in terms of the fluorescence fold change ($\Delta F/F_0$). For the concentration series of S RBD analyte tested (**Figure 2D-E**), the *LOB* is calculated to be $\Delta F/F_0$ = 0.2828. The limit of detection is then:

$$LOD = LOB + 1.645\ SE_{low\ conc.}$$

Where the low concentration is set as the fluorescence values corresponding to 50 nM S RBD. Thus, the *LOD* is calculated to be $\Delta F/F_0$ = 0.2919, which corresponds to a S RBD concentration of 9.49 nM based on the cooperative binding model fit.

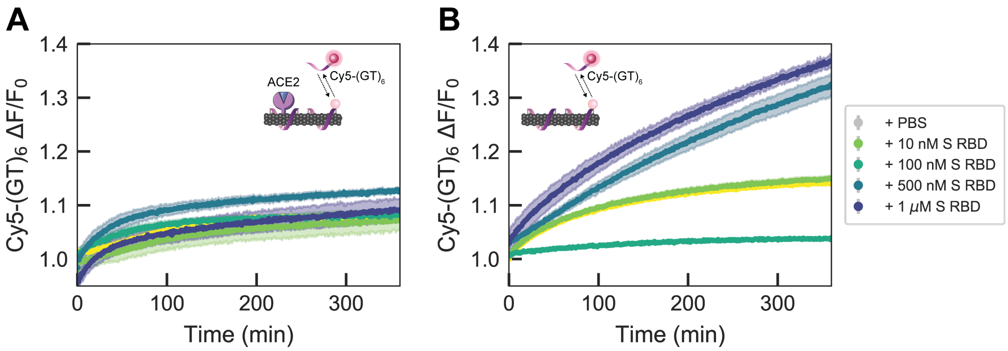


**Figure S8. Displacement of Cy5-(GT)$_6$ ssDNA from SWCNT in the presence or absence of ACE2 sensing protein, as a function of S RBD analyte concentration.** Addition of varying concentrations of S RBD analyte to **(A)** ACE2-Cy5-(GT)$_6$-SWCNT nanosensors (formed by 6.25 mg/L ACE2 and 2.5 mg/L (GT)$_6$-SWCNTs) and **(B)** Cy5-(GT)$_6$-SWCNTs alone (2.5 mg/L). Shaded error bars represent standard error between experimental replicates (N = 3).

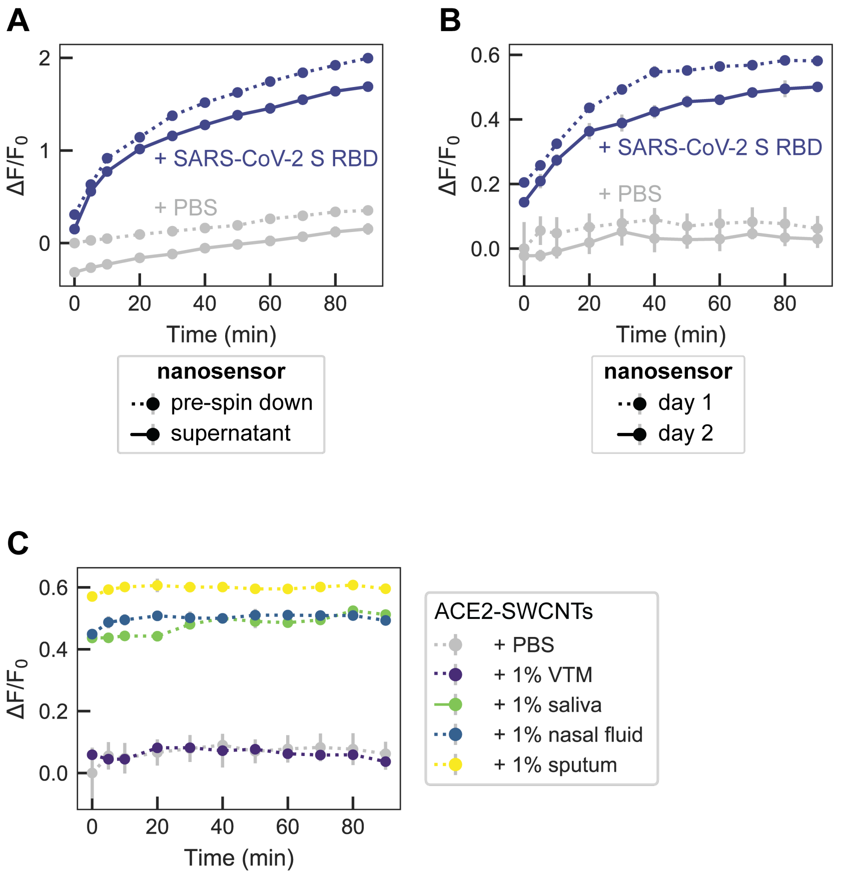


**Figure S9. ACE2-SWCNT nanosensor controls for assessing stability.** Response of ACE2-SWCNTs (formed by 6.25 mg/L ACE2 and 2.5 mg/L $(GT)_6$-SWCNTs) to S RBD was preserved **(A)** before and after centrifugation (16.1 krcf, 30 min; 500 nM S RBD) and **(B)** before and after overnight incubation at ambient conditions (1 μM S RBD). **(C)** Stability of ACE2-SWCNT nanosensors in different biofluids. Normalized change in fluorescence of the 1130 nm SWCNT emission peak for the ACE2-SWCNT sensor as a function of time in 1% relevant biofluids: viral transport medium (VTM), saliva, nasal fluid, and sputum (treated with sputasol). Nanosensor fluorescence in biofluids demonstrated stability yet elevated magnitudes.

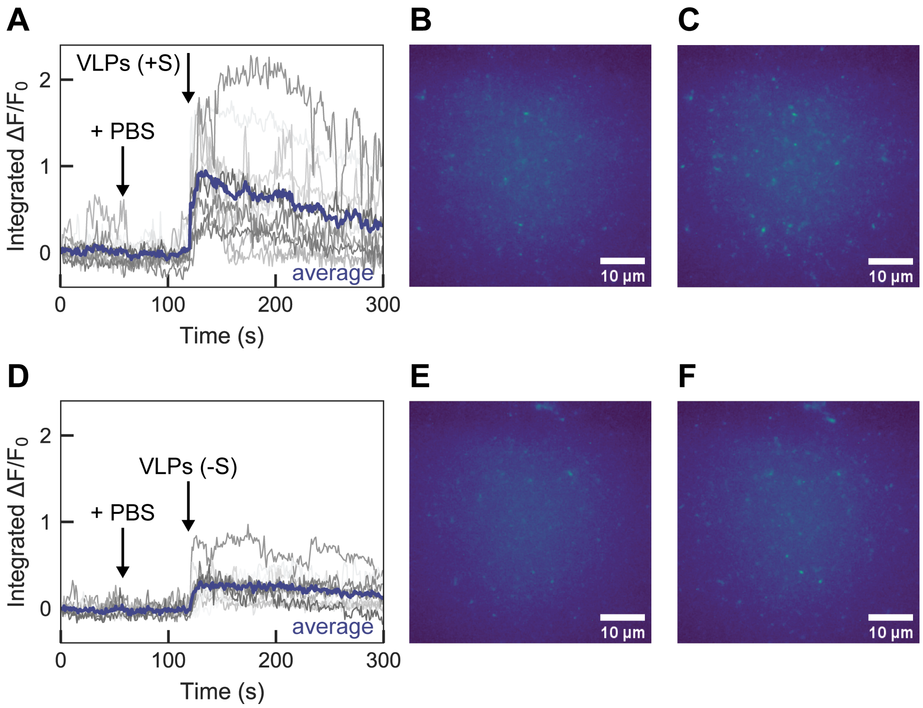


**Figure S10. Surface-immobilized ACE2-SWCNT nanosensor response to virus-like particles (VLPs) with and without S protein.** Single-molecule microscopy traces of ACE2-SWCNTs (formed by 12.5 mg/L ACE2 and 5 mg/L $(GT)_6$-SWCNTs) immobilized on a glass microwell dish exhibited a larger fluorescence response to VLPs expressing S protein, for single regions of interest (gray; 12 total per image) and the average intensity (purple). **(A-C)** Addition of PBS at 60 s caused no change in fluorescence, as expected, and addition of 50 mg/L VLPs (no S protein) at 120 s yielded a minor turn-on fluorescence response, as shown by **(A)** the integrated-fluorescence fold change ($\Delta F/F_0$) over 5 min and entire field-of-view at **(B)** time = 100 s and **(C)** time = 125 s. **(D-F)** Addition of PBS at 60 s caused no change in fluorescence, as expected, and addition of 50 mg/L VLPs (with S protein) at 120 s yielded a large turn-on fluorescence response, as shown by **(D)** the integrated-fluorescence fold change ($\Delta F/F_0$) over 5 min and entire field-of-view at **(E)** time = 100 s and **(F)** time = 125 s. All fluorescence images were obtained with 721 nm laser excitation and a 100x oil immersion objective.

This platform for nanosensor design was expanded to attempt passivation of $(GT)_6$-SWCNTs with an antibody for SARS-CoV-1 S RBD that has been previously verified to bind CoV-2 S RBD (*5*). Interestingly, the anti-S sensing protein did not exhibit any modulation in intrinsic SWCNT fluorescence, nor did addition of the S RBD analyte provoke a fluorescence response. This result implying a lack of interaction between anti-S and the SWCNT surface is in line with our previous work that implies antibodies exhibit minimal adsorption to ssDNA-SWCNTs (*6*). Thus, alternative attachment strategies must be pursued for sensing proteins with no intrinsic affinity for the SWCNT surface.

**Table S1.** Purchased biofluid and protein specifications.

| Purpose | Protein | Manufacturer | Lot # | Source | Form | Concentration |
|---|---|---|---|---|---|---|
| Sensing protein | Angiotensin-converting enzyme 2 (ACE2) | Ray Biotech | 04U24020GC | Recombinant, (HEK293 cell expression system; C-terminal His-tag) | Liquid (PBS) | 2.8 g/L |
| Control | SARS CoV-1 spike protein receptor-binding domain (CoV-1 S RBD) | ACROBiosystems | 3558c-203KF1-R7 | Recombinant (HEK293 cell expression system; C-terminal His-tag) | Lyophilized from PBS, 10% Trehalose | Reconstituted in 167 μL 0.1 M PBS, 30 min at room temperature with occasional gentle mixing |
| Control | MERS spike protein receptor-binding domain (MERS S RBD) | MyBioSource | 0110YB | Recombinant (HEK293 cell expression system; C-terminal His-tag) | Liquid (PBS, 0.1% sodium azide) | 1 g/L |
| Control | Influenza hemagglutinin subunit (FLU HA1) | MyBioSource | 95-101-1104 | Recombinant (E. coli expression; N-terminal His-tag and strepII-tag) | Liquid (PBS, 0.1% SDS, 0.02% sodium azide) | 1 g/L |
| Control | Human serum albumin (HSA) | Sigma-Aldrich | #SLBZ2785 | Human plasma | Lyophilized, fatty acid and globulin free | Reconstituted in 0.1 M PBS, 30 min at room temperature with occasional gentle mixing |
| Biofluid | Viral transport medium | Innovative Research | 31975 | N/A | Liquid | N/A |
| Biofluid | Saliva | Lee Biosolutions | W205820 | Human, pooled, normal | Liquid | N/A |
| Biofluid | Nasal fluid | Lee Biosolutions | 18-05-538 | Human, single donor, normal | Liquid | N/A |
| Biofluid | Sputum | Lee Biosolutions | 18-03-594 | Human, single donor, normal | Liquid | N/A |
